# Tricuspid annulus remodelling after transcatheter edge-to-edge repair for reduction of tricuspid regurgitation

**DOI:** 10.1101/2023.07.24.23293107

**Authors:** Maddalena Widmann, Fausto Castriota, Roberto Nerla, Flavio Ribichini, Gabriele Pesarini, Andrea Fisicaro, Angelo Squeri

## Abstract

**Background:** In the last years multiple transcatheter devices for tricuspid valve interventions were developed. Aim of the study is to evaluate the acute tricuspid annulus remodelling after percutaneous leaflet repair, using a leaflet approximation device (TriClip Abbott Vascular, Santa Clara, CA, USA) for reduction of tricuspid regurgitation.

**Methods and results:** This is a retrospective dual-center cohort study that includes 26 consecutive patients, treated with TriClip in Maria Cecilia Hospital of Cotignola and in the University Hospital of Verona. Tricuspid annulus geometry was evaluated using three-dimensional transesophageal echocardiography examinations conducted during the procedure before and after TriClip implantation.

The mean age of the study cohort was 79,3 years, and 88,5% were female. Tricuspid regurgitation was graded severe or greater at pre-operative examination in all patients, mostly due to annular dilation. Procedure was successfully in all patients, with at least 1-grade reduction of tricuspid regurgitation before hospital dismissal. A significative reduction of mean septal-lateral diameter (4,09 ± 0,44 cm vs 3,54 ± 0,53 cm, p=< 0,0001), mean major diameter (4,65 ± 0,63 cm vs 4,28 ± 0,65 cm, p=0,0002), planimetric area (14,00 ± 2,91 cm2 vs 11,25 ± 2,91 cm2, p=<0,0001) and perimeter (13,62 ± 1,43 cm vs 12,42 ± 1,62 cm, p=<0,0001) of the tricuspid annulus was observed.

**Conclusions:** In this small real-world population, edge-to-edge repair using TriClip was found to be effective and safe. Tricuspid transcatheter repair with a leaflet approximation device lead also to reduction in the tricuspid annular dimensions. This is to date the first study that shows positive changes in tricuspid annular geometry, that could have potentially relevant therapeutic implications.

## Introduction

Clinically relevant tricuspid regurgitation (TR) is a prevalent valvular disease that interests 3,0 million individuals in Europe and 1,6 million in the USA.^1^ TR is classified according to the underlying mechanism in primary, secondary and isolated. Severe TR has a significant impact on survival and is associated with worsening heart failure. ^1^ ^2^ A retrospective analysis of a large cohort of patients shows one-year mortality rates of 29,5% for moderate TR and 45,6% for severe TR, independently of potential confounders. Also the rate of heart failure hospitalization is positively correlated with TR severity. ^3^

Transthoracic echo plays a key role in the diagnosis of TR, in order to quantify TR severity, distinguish primary from secondary or isolated TR, define any associated left-sided disease and evaluating right ventricle (RV) size and function. ^4^ The grading of TR, traditionally definite as mild, moderate and severe, requires a comprehensive, multi-parametric approach. To better characterize the severity of TR, a new grading scheme including two additional grades “massive” and “torrential” has been proposed.^5^ This scheme is now used in clinical studies on transcatheter interventions to standardize and quantify treatment effects. Indeed, some studies demonstrated that a reduction of TR of a full grade or more is clinically meaningful and associated to an improvement in quality of life.^6^

Tricuspid valve (TV) has a complex anatomical structure, that comprehends annulus, papillary muscle, leaflets, and chordae and the interdependence between TV and RV explains the physiopatological mechanisms of most forms of TR. The tricuspid annulus (TA) is a nonplanar structure, with a more ventricular posteroseptal portion and a more atrial anteroseptal portion, near to the RV outflow tract and the aortic valve. It is also a dynamic structure that varies his shape and size according with loading conditions and throughout the cardiac cycle. A study demonstrated that in patients with functional TR, TA has a more circular shape resulting from dilation in the septal to lateral and posteroseptal to anterolateral directions. Moreover, the TA becomes more planar, the more severe is the TR. ^7^

Despite TR being markedly undertreated, a better understanding of the pathophysiology and the prognostic implications of TR have recently shifted the treatment from a conservative to a more interventional approach. The disease is surgically treated mostly using various plasty techniques targeting annular dilatation, but the appropriate timing of intervention is still largely debated. In particular, there are no recommendations for patients with severe isolated TR and surgery is seldom performed in this setting due to high operative mortality. For this reason, multiple transcatheter devices for TV interventions are currently in clinical trials and are considered valuable treatment options in anatomically eligible patient at high surgical risk. Percutaneous devices involve similar techniques as surgical repair and a recent analysis shows significant improvement of quality of life with fewer periprocedural complications. ^8^ Efficacy and clinical outcome are promising ^9 10^, but the effect on RV reverse remodelling is still debated.

Percutaneous technique currently available are derived from surgery and are the leaflet-approximation technique that aim to reduce TR through leaflet grasping and the transcatheter annuloplasty. The most studied device for percutaneous annuloplasty is the Cardioband™ (Edwards Life-sciences, Irvine, CA, USA) system that addresses the pathophysiological mechanism of annular dilatation and consists of the implantation of an adjustable band on the TA using several screws as anchors. The TriClip (Abbott Vascular, Santa Clara, CA, USA) and the PASCAL (Edwards Lifescience, Irvine, CA,USA) are transcatheter leaflet repair system that act approximating tricuspid leaflets and in a propensity matched analysis their outcomes are comparable. ^11^The TriClip system consist of a guiding catheter and a clip delivery system that are designed to navigate in the RV. The last generation of the device has longer and wider arms allowing for more efficient grasping. ^12^ Analysis in the real word setting shows an high procedural success rate with reduction in TR of at least one grade in most of patients. ^13 14^ RV remodelling after percutaneous TV interventions has to date few evidences, but the available data are promising. ^15 9^ Analysis of the hemodynamic consequences of transcatheter tricuspid edge-to-edge repair in patient with isolated TR shows that reduction of regurgitant volume leads to a decrease of RV volume overload with reduction in RV diastolic dimensions and total RV stroke volume.^16^ In the other hand, this causes an augmentation of pulmonary forward flow with subsequent enhanced left ventricle filling and improvements in cardiac index. Also a reduction in external work of the RV is observed, with improvement of RV performance and reduction of oxygen comsumption.^17^

## Methods

### Aim of the study

The study evaluates the acute TA remodelling after percutaneous leaflet repair. One of the most important mechanisms of secondary or isolated TR is the annular dilatation but TriClip procedure target are the tricuspid leaflets. Authors wanted to test if leaflet approximation technique can also lead to a reverse TA remodelling as a potential mechanism for TR reduction.

### Study population

This is a retrospective dual-center cohort study that includes 36 consecutive patients, treated with transcatheter edge-to-edge repair for reduction of tricuspid regurgitation using TriClip. From December 2017 to February 2023, 34 patients were treated in Maria Cecilia Hospital of Cotignola and 2 in the University Hospital of Verona. Transthoracic and transesophageal examinations were analysed and 10 patients without suitable intraoperative TEE 3D reconstruction of the TV before and after TriClip implantation were excluded For each patient demographic variables, cardiovascular risk factors and blood samples were collected at admission (Table 1). Echocardiographic parameters at the pre-operative and post-operative transthoracic and transesophageal examinations were measured (Table 2). All echocardiograms were performed on a IE33 or EpiqCVX (Philips Medical System, Andover MA) according to current guidelines by the European Association of Cardiovascular Imaging and American Society of Echocardiography. Quantification of TR is made integrating qualitative and quantitative approach and is described using five grades. The TEE 3D examinations used for evaluation of TA were conducted during the procedure after induction of general anaesthesia, before and after Triclip implant. The reconstructions were analysed using QLab (Philips Medical Systems, Andover, MA) software package from stored images by an experienced operator blinded to procedural details. Procedural data were also collected (Table 3).

**Table 1.**
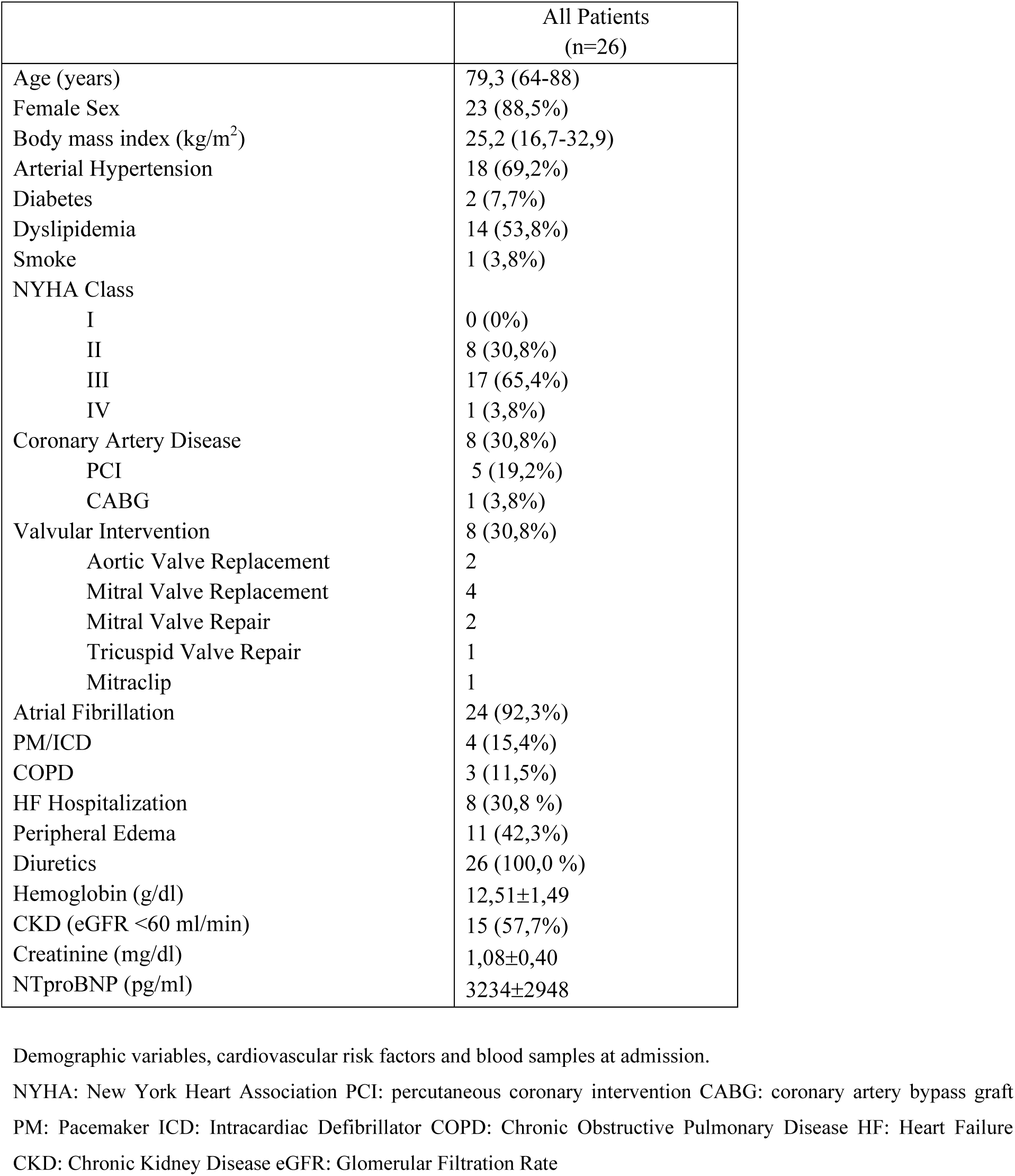

**Table 2.**
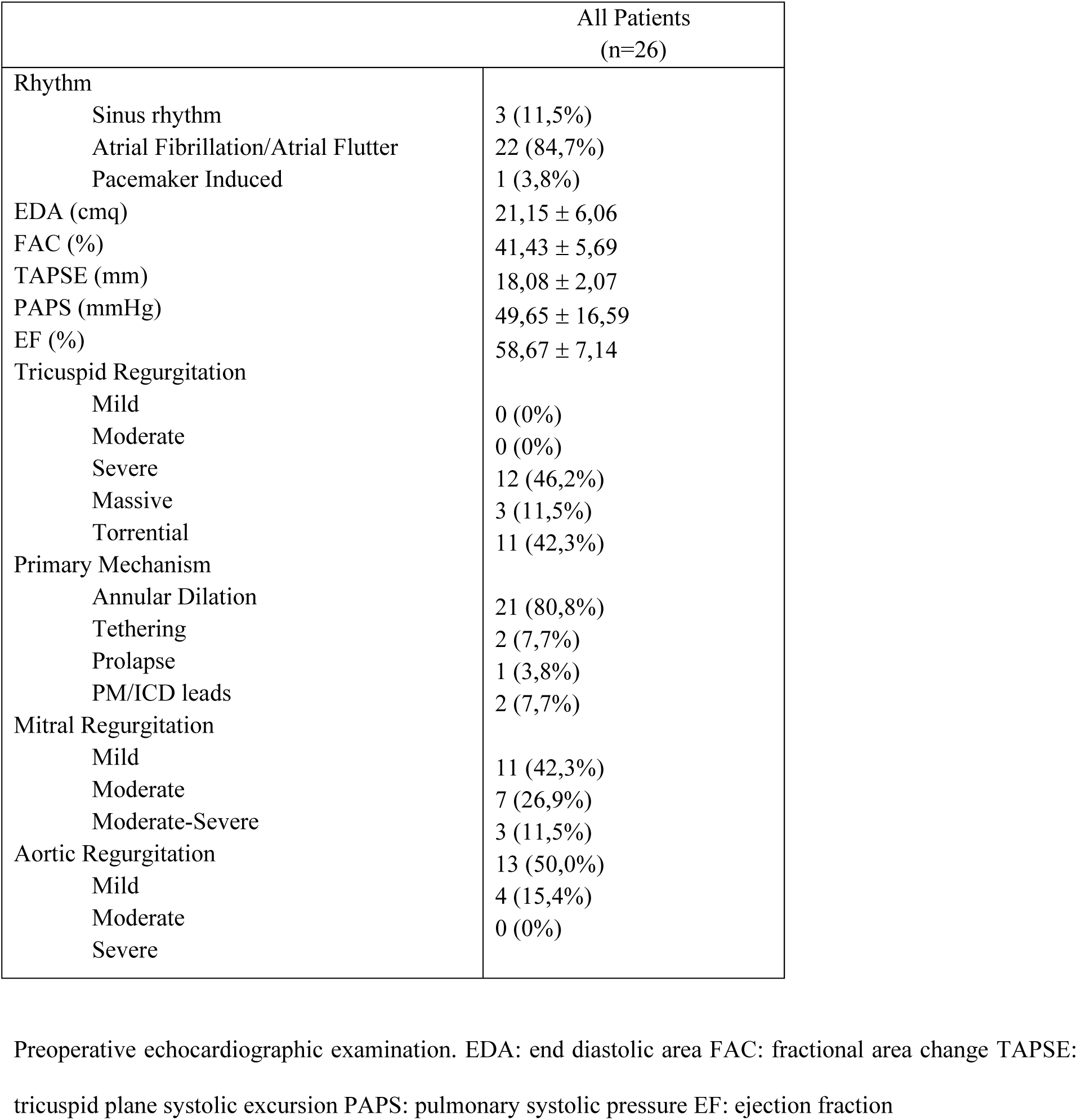

**Table 3:**
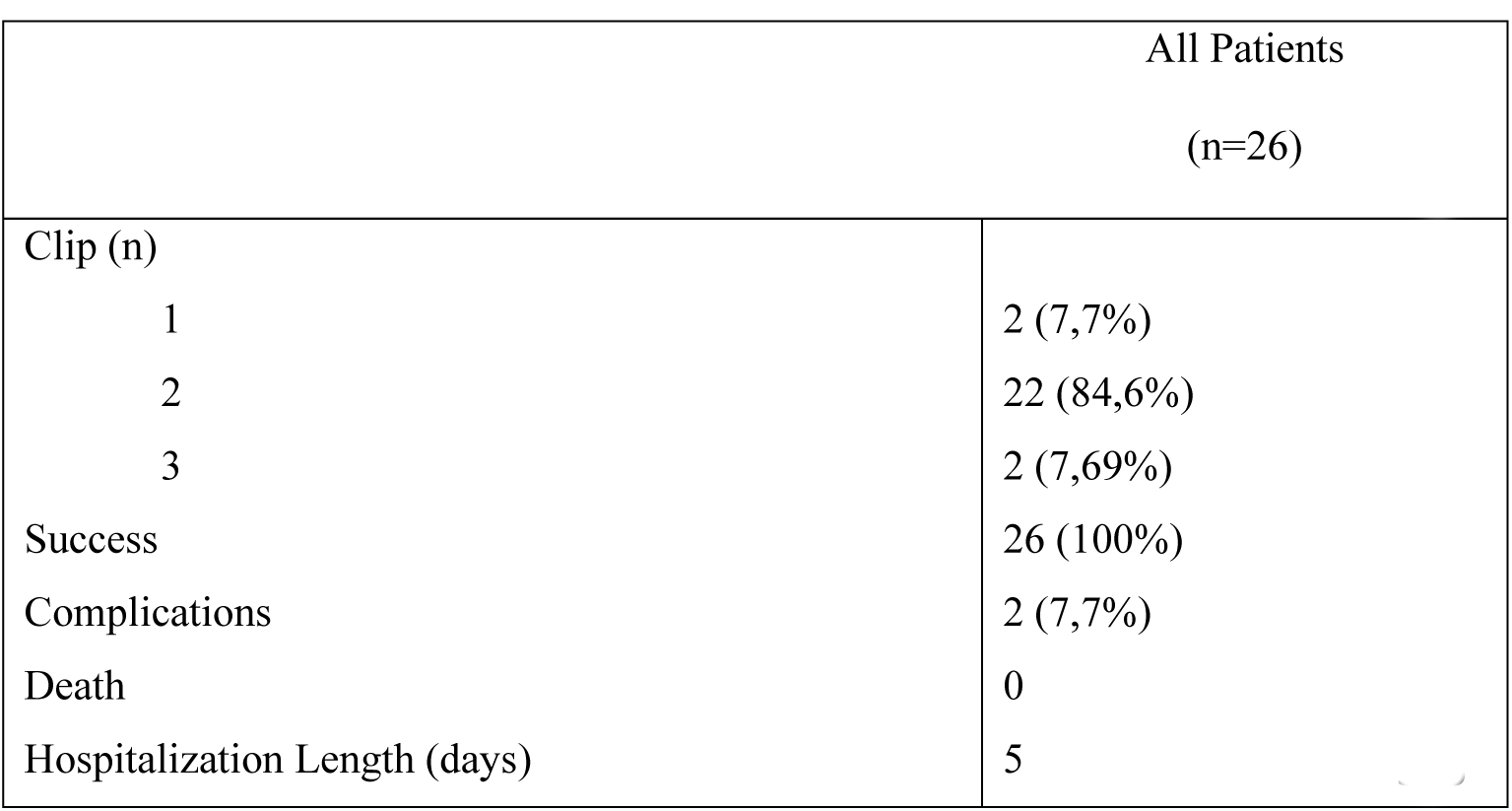
Operative Outcomes.

The study was conducted in accordance with the ethical principles of the Declaration of Helsinki and all the patients included entered in the prospective clinical registry and provided their written consent for the anonymous collection of the data.

### Tricuspid annulus analysis

The QLab software was used to analyse TA from intraprocedural 3D examinations. Manipulation of the 3D data sets by MPR was performed to accurately identify the maximal septal-lateral (SL) and antero-posterior (AP) diameters and to calculate perimeter and area of the TA. Also, the major diameter of the planimetric TA was measured. Analysis was conducted at the end-diastole. The SL and AP dimensions were measured respectively in the coronal and the sagittal plane, using some landmarks as the aortic root for correct alignment. The TA was planimetered in the transverse plane to obtain perimeter, area, and major diameter. (Figure 1) For all patients, the same analysis is performed from data sets collected before and after TV repair, using the same method. The eccentricity index was obtained dividing SL diameter by AP diameter.

**Figure 1.**
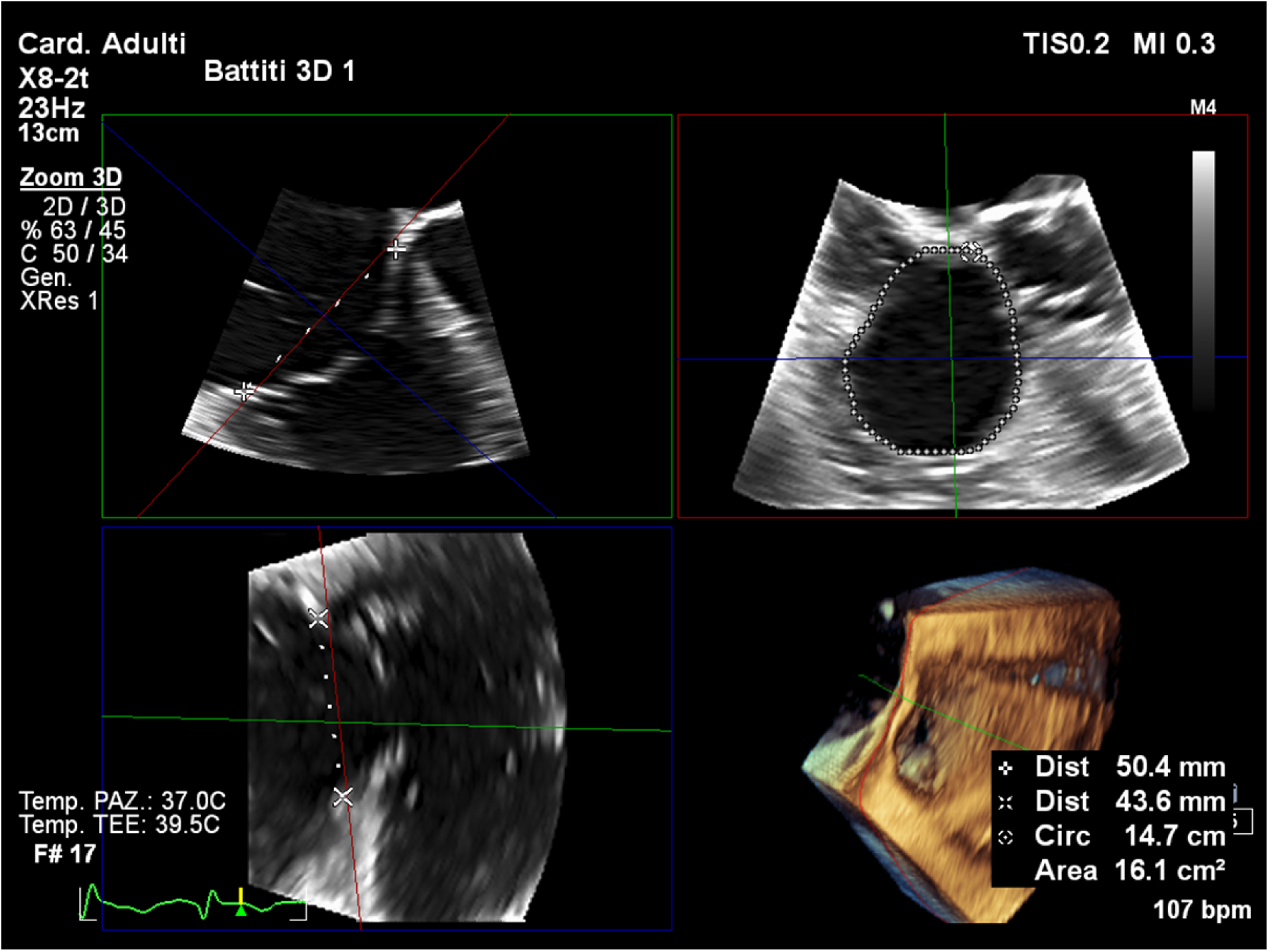
Tricuspid annular measurement using multi-planar-reconstruction.

### Statistical Analysis

For descriptive analyses, categorical variables are presented as numbers and percentages and continuous variables are presented as means ± standard deviation. The parameters of AP, SL, major diameter as well area and perimeter of the TA were compared before and after TriClip implantation using t-test for paired data. Eccentricity index was also compared using the same statistical method.

## Results

### Clinical and echocardiographic characteristics

The mean age of the study cohort was 79,3 years, and 88,5% were female. Severe functional limitation (New York Heart Association ≥ III) was seen in 69,2% of patients. 8 patients had prior valve intervention and atrial fibrillation (AF) was identified preoperatively in 92,3% (n = 24) of patients. 4 patients had prior PM/ICD implantation and 8 had coronary artery disease. At admission, a significant proportion of patients (42,3% n=11) had peripheral oedema and all patients were on diuretic therapy. 8 (30,8%) had one or more previous hospitalization for heart failure and mean value of NT-proBNP was 3234 pg/ml. At pre-operative transesophageal and transthoracic echocardiographic examination, RV systolic dysfunction was identified in 3 patients (11,5%), with mean fractional area change of 41,43 ± 5,69% and mean tricuspid plane systolic excursion of 18,08 ± 2,07 mm. The mean pulmonary systolic pressure was 49,65 ± 16,59 mmHg. All patients had preserved left ventricular ejection fraction and 3 (11,5 %) of them had moderate-severe mitral regurgitation. (Table 2). TR was graded severe or greater at pre-operative examination in all patients, mostly due annular dilation(Table 2).

### Operative Outcomes

Edge-to-edge reparation was successfully in 100% (n=26) with at least 1-grade reduction of TR at the end of the procedure (Figure 2). Most patients (84,6% n=22) implanted two clips. New permanent pacemaker implantation was required in 1 patient for a second-degree atrioventricular block and another patient had a vascular complication of the operative access. The median length of a hospital stay was 5 days. (Table 3)

**Figure 2.**
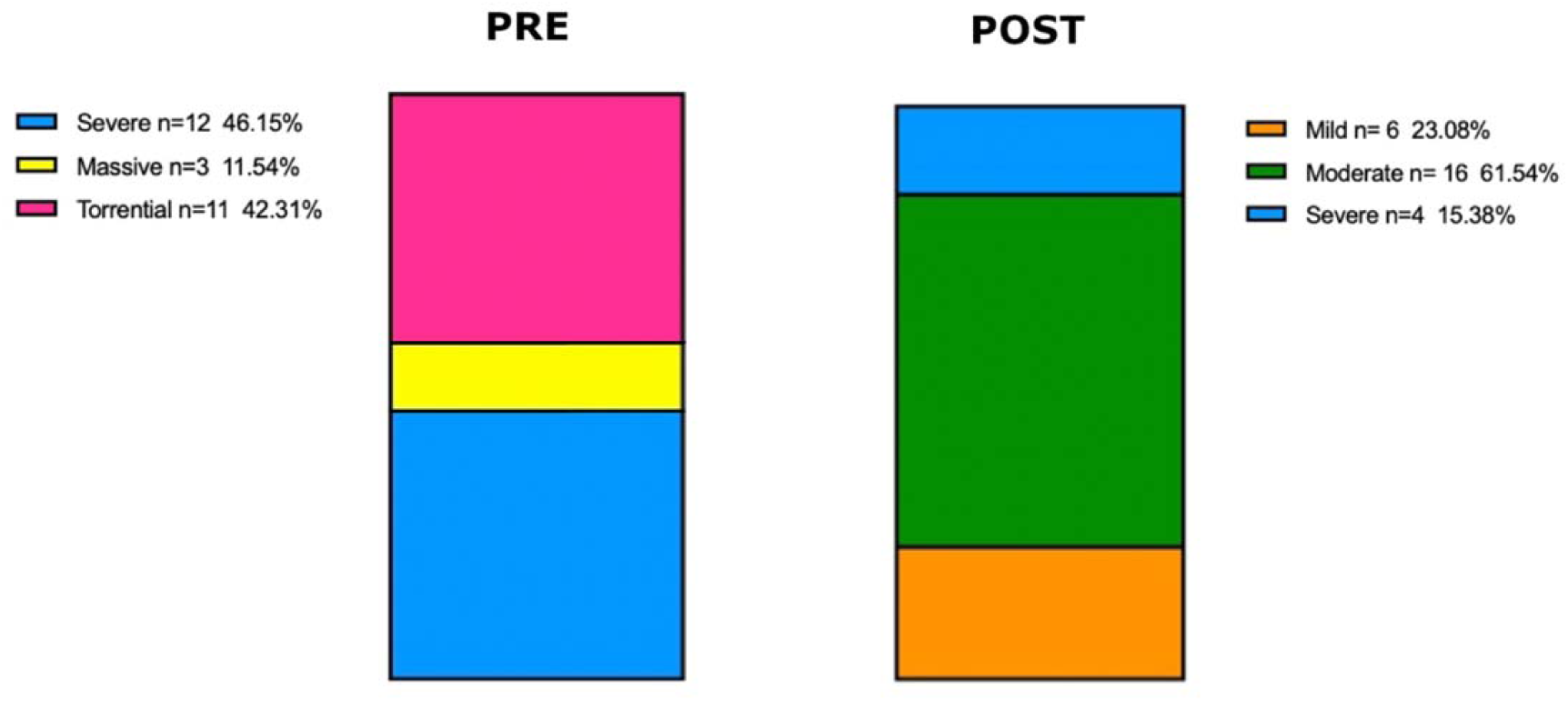
Tricuspid regurgitation quantification pre- and post-intervention.

### Echocardiographic outcomes

Before hospital dismissal 23,1% (n=6) of patients had mild TR (Table 4). TR grade was reduced to at least moderate in all but 2 (7,69%) patients. All (100% n=26) had at least 1-grade reduction of TR. (Figure 5) Mean tricuspid plane systolic excursion was 17,29 ± 3,58 and mean left ventricle ejection fraction 59,96 ± 8,83%.

**Table 4:**
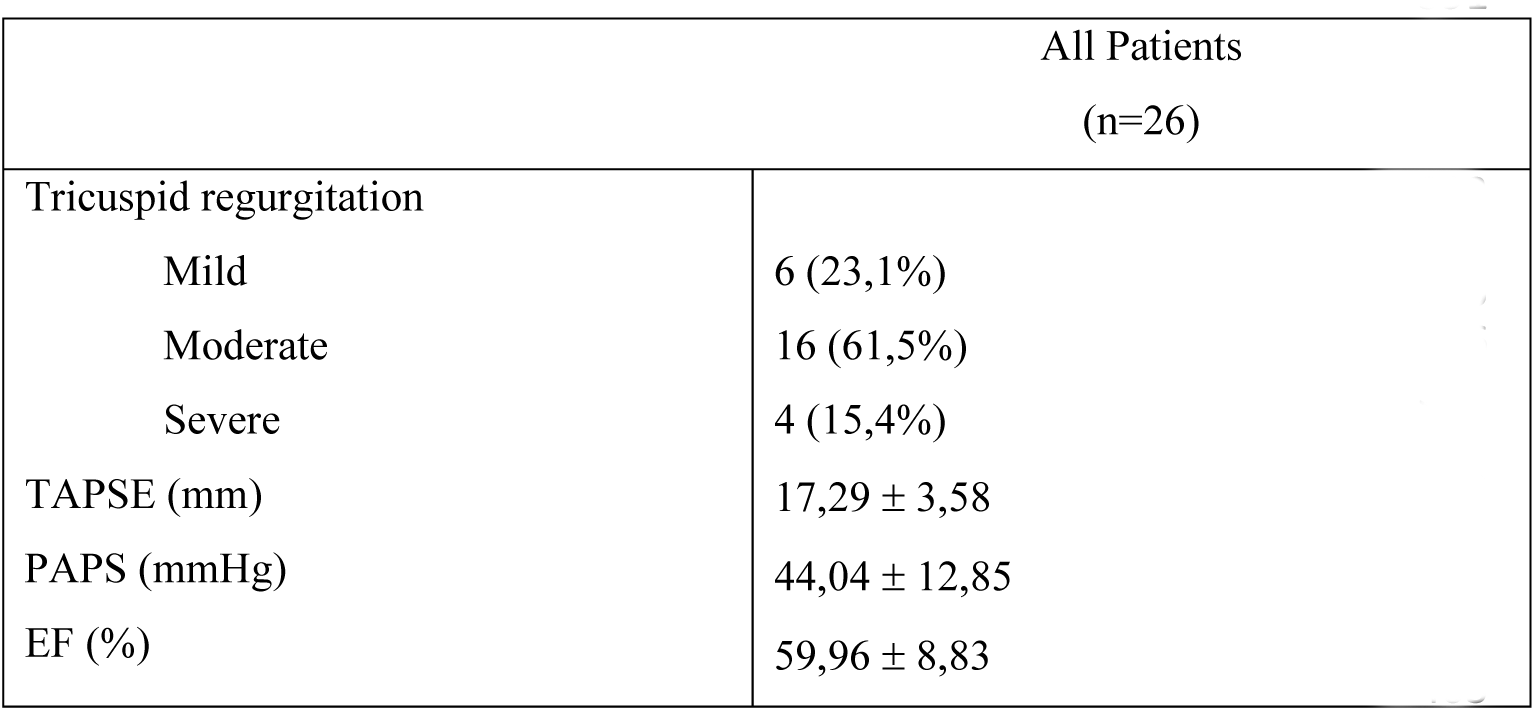
Echocardiographic outcomes.

Echocardiographic evaluation at follow-up, 1 month after procedure, didn’t show any difference in terms of tricuspid regurgitation.

### Acute tricuspid annulus remodelling

Analysis of the TA dimension before and after TriClip implantation (Table 5) showed a significative reduction of the mean SL diameter (p=< 0,0001) and of the mean major diameter (p=0,0002), whereas the reduction of the AP diameter didn’t reach the statistical significance (p=0,0626). Comparing the planimetric area and perimeter of the annulus before and after the procedure (Figure 3) a significative reduction was observed (p=<0,0001, p=<0,0001). Finally, the mean eccentricity index was 0,98 ± 0,17 before TriClip implantation and 0,90 ± 0,17 after (p=0,0286).

**Figure 3.**
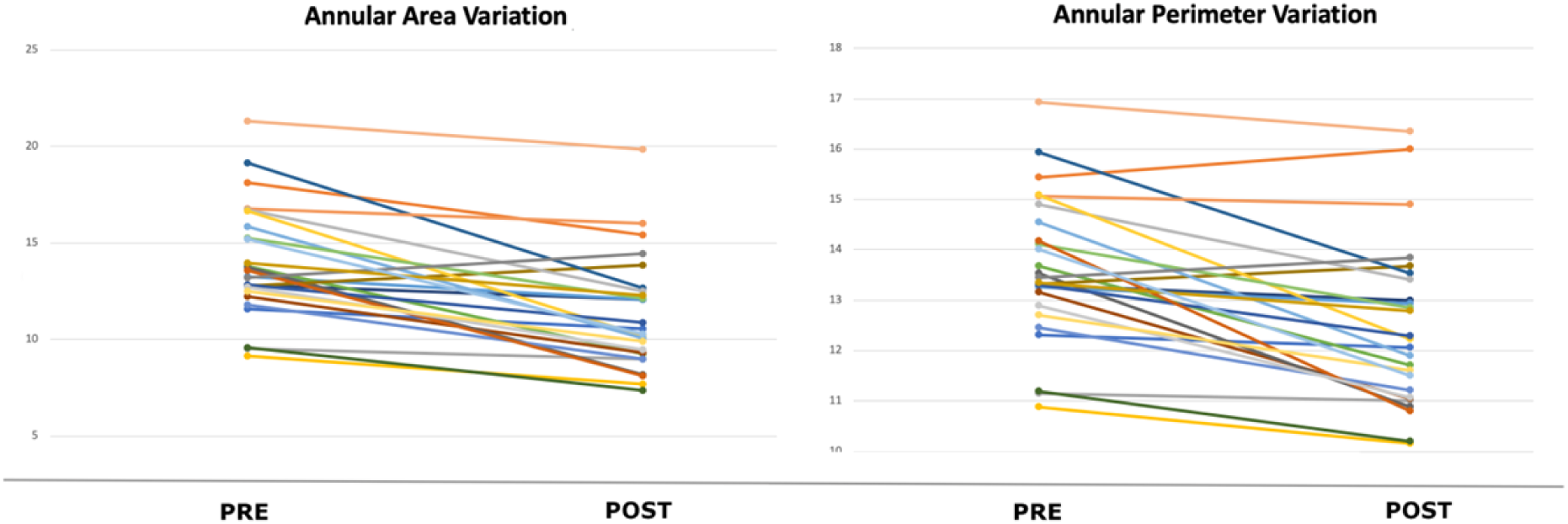
Variation in tricuspid annular area and perimeter pre- and post-procedure in every single patients.

**Table 5:** Acute tricuspid annulus remodelling.

|  | Before | After | P value |
| --- | --- | --- | --- |
| SL diameter (cm) | 4,09 ± 0,44 | 3,54 ± 0,53 | < 0,0001 |
| AP diameter (cm) | 4,29 ± 0,79 | 4,01 ± 0,62 | 0,0626 |
| Major diameter (cm) | 4,65 ± 0,63 | 4,28 ± 0,65 | 0,0002 |
| Area (cmq) | 14,00 ± 2,91 | 11,25 ± 2,91 | < 0,0001 |
| Perimeter (cm) | 13,62 ± 1,43 | 12,42 ± 1,62 | <0,0001 |
| Eccentricity Index | 0,98 ± 0,17 | 0,90 ± 0,17 | 0,0286 |

SL: septal-lateral diameter AP: antero-posterior diameter

## Discussion

In this retrospective, dual-center cohort study, TA dimensions were analysed before and early after edge-to-edge tricuspid leaflet repair using TriClip in a real-world population. The study population is composed of older people, mostly women with isolated functional TR. In fact, only few patients have associated left-sided valvular or ischemic disease, and no-one has left ventricular systolic dysfunction. However, almost all patients have anamnestic AF and so the predominant mechanism of TR in the population is annular dilation and right atrial remodelling secondary to AF. The study population is comparable with those of the pivotal studies of the TriClip and PASCAL systems and because of the prohibitive surgical risk due to age, comorbidities and isolated tricuspid valve disease this patients are good candidates for percutaneous therapies. ^18 19^

The study demonstrates a procedural success in all patients, with correct implantation of one or more devices. No life-threatening complication are observed, confirming a good safety profile of the system. A five-class grading scheme is used in the pre-operative echocardiographic transthoracic and transesophageal evaluation to assess the severity of TR. Before dismissal, all patients showed a reduction of TR of at least 1-grade and only four of them had severe residual TR. In addition, no patient developed acute right ventricular dysfunction.

TR caused by annular dilation has been surgically treated for decades by ring annuloplasty. To date, companies are evaluating some percutaneous systems with the aim to directly reduce annular size similarly to those surgical rings. Also the edge-to-edge percutaneous systems are derived from surgery, ^20^ that initially developed this techniques for treatment of TR due to complex lesions not treatable with annuloplasty only. TriClip and the other available edge-to-edge reparation systems reduce the valve regurgitation stitching together the tricuspid leaflets and limiting their motion. The reduction of volume overload generates a positive reverse remodelling of the RV but the indirect action on TA has not been to date fully elucidated. This observational study sought to evaluate the acute remodelling of the TA, after TriClip implant. The retrospective analysis of 3D echocardiographic intraprocedural evaluations demonstrated that edge-to-edge tricuspid reparation using TriClip determinates in the study population a positive acute remodelling of the TA, with significative reduction of the SL and the major diameter. Also, planimetric area and perimeter decreased significantly, corroborating the echocardiographic outcomes. The analysis of the AP diameter highlights a reduction that does not reach the statistical significance and this result could be associated to the evidence of decreasing of the eccentricity index. In fact, study demonstrated that TA dilates mostly at the lateral region, because the septal region is more rigid and fibrous. For this reason, the coaptation gap is often located between anterior and septal leaflet and most of the clips were placed in the antero-septal coaptation line (Table 6). It could have twisted the TA in some patients, giving it a more oval shape and decreasing his eccentricity. Our data suggest that both, acute RV volume reduction and leaflefts approximation forced by the device itself, lead to an annular positive remodelling. Tricuspide annular dimension reduction is on the other hand one of the mechanism of TR severity reduction.

**Table 6:** Position of the implanted clips.

|  |  |
| --- | --- |
| A-S | 82,9% |
| P-S | 12,2% |
| A-P-S | 4,9% |
| A-P | 0% |

A=anterior leaflet S=septal leaflet P=posterior leaflet

Previous reports suggest that the mechanism of TR related mortality is progressive RV dysfunction and fibrosis. We hypothesize that the reduction of volume overload associated with positive remodelling of TA could reduce symptoms and positively effect on prognosis interrupting this vicious cycle. Follow-up data with clinical evaluation and analysis of the TA are needed to confirm this thesis.

### Limitations

Several limitations should be acknowledged. First, this study was performed retrospectively, based on a small sample size cohort without any control subject. Therefore, these results need to be confirmed in a larger, prospective study. Furthermore, the quality of intraprocedural imaging can be suboptimal because of shadowing by intracardiac implants or by TriClip device itself and this aspect could affect the analysis with the reconstruction software. In addition, no prognostic implications were provided.

## Conclusion

In this small real-world population, edge-to-edge repair using TriClip was found to be effective and safe. This is to date the first study that shows a positive acute remodelling of the TA after TriClip implantation suggesting that the efficacy of TR reduction is not due to leaflet approximation itself but by annular reduction too.

### Sources of Funding

The authors declare that no funds, grants, or other support were received during the preparation of this manuscript.

### Disclosures

None.

### Clinical Perspective

Clinical Competencies in “Advanced Sub Speciality Cardiovascular training: Echocardiography”

The effect of leaflet approximation on tricuspid annular remodelling has several phisiopatological implication. Since annular dilatation is the most important mechanism in the development of tricuspid regurgitation the demonstration of annular dimension reduction after Triclip procedure give an explanation on the efficacy of leaflets approximation therapy over the forced coaptation effect.

As a future scenario, the amount of annular reduction after Triclip could be a surrogate of long term results in terms of freedom from tricuspid regurgitation recurrence at follow-up. These could be investigated in a more wide population and with a prolonged follow-up in order to assess the relationship between annular reduction ad recurrence of tricuspid regurgitation.

## Abbreviations

TR: tricuspid regurgitation
TA: tricuspid annulus
RV: right ventricle
TV: tricuspid valve
3D: three-dimensional
CMR: cardiac magnetic resonance
TEE: transesophageal echocardiogram
AF: atrial fibrillation
MPR: Multi Planar Reconstruction

## Data Availability

All data produced in the present work are contained in the manuscript.

